# Restoration of normal central pain processing following manual therapy in nonspecific chronic neck pain

**DOI:** 10.1101/2023.10.26.23297616

**Authors:** Josu Zabala Mata, Jon Jatsu Azkue, Joel E. Bialosky, Marc Saez, Estíbaliz Dominguez López, Maialen Araolaza Arrieta, Ion Lascurain-Aguirrebeña

**Author notes:** Corresponding author (JZ). These authors contributed equally to this work (Josu Zabala Mata, Jon Jatsu Azkue, Joel E. Bialosky, Ion Lascurain-Aguirrebeña). These authors also contributed equally to this work (Marc Saez, Estíbaliz Dominguez López, Maialen Araolaza Arrieta).

## Abstract

**Objective:** to determine if a 4-week manual therapy treatment restores normal functioning of central pain processing mechanisms in non-specific chronic neck pain (NSCNP), as well as the existence of a possible relationship between changes in pain processing mechanisms and clinical outcome.

**Design:** cohort study.

**Methods:** sixty-three patients with NSCNP received four treatment sessions (once a week) of manual therapy. Pressure pain thresholds (PPTs), conditioned pain modulation (CPM) and temporal summation of pain (TSP) were evaluated at baseline and after treatment completion. Therapy outcome was measured using the Global Rating of Change Scale, the Neck disability Index, intensity of pain during the last 24 hours, Tampa Scale of Kinesiophobia and Pain Catastrophizing Scale.

**Results:** Following treatment, an increased CPM response and attenuated TSP were found, along with amelioration of pain and improved clinical status. PPTs at trapezius muscle on the side of neck pain were increased after therapy, but not those on the contralateral trapezius and tibialis anterior muscles. Only minor associations were found between normalization of TSP/CPM and measures of clinical outcome.

**Conclusion:** Clinical improvement after manual therapy is accompanied by restoration of CPM and TSP responses to normal levels in NSCNP patients. The existence of only minor associations between changes in central pain processing and clinical outcome suggests multiple mechanisms of action of manual therapy in NSCNP.

## Introduction

Neck pain is among the top five causes of Disability in middle- and high-income countries and among the top ten as a cause of global disability(1). Despite investment in research, the prevalence of neck pain has not declined substantially in the last two decades(2). Moreover, recurrence reaches 50-75% within the next 5 years following the first episode,(3,4) and 68% of individuals experiencing an episode of acute neck pain will become chronic neck pain sufferers(5). Since little relationship with radiological findings and no specific cause is found to explain symptoms, they are usually classified as patients with non-specific neck pain (NSCNP)(6).

Guidelines advocate treating patients with NSCNP with exercise and manual therapy(7). Despite the widespread use of manual therapy, systematic reviews assessing clinical outcomes report low to moderate treatment effects at best(8). The lack of larger reported effects should be of no surprise, since several aspects of treatment remain to be established, such as optimal dosage and clinical parameters, best indicated forms of mobilization, and possible target patient subpopulations. This may be partly due to the fact that mechanisms of action of manual therapy are not yet fully understood. Although biomechanical effects(9), neural hysteresis,(10) and segmental neurological modulation(11) have long been postulated as underlying mechanisms of action of manual therapy, hypotheses have in recent years shifted towards a potential role of central nervous system pain processing(12).

Inter-individual variability in the functioning of central pain processing mechanisms has been postulated as an alternative framework to understand heterogeneity of treatment outcomes(13). Several studies have reported disturbances in central pain processing in patients with NSCNP(14–16). A meta-analysis has confirmed the occurrence of hyperalgesia distal to the most painful site, a probable indication of the occurrence of central sensitization in the NSCNP population^19^. The phenomenon of central sensitization (CS) is a state of increased central responsiveness to nociceptive inputs associated with plastic changes in nociceptive circuits and pathways(17). There is consistent evidence of altered central pain processing in patients with NSCNP, including both pronociceptive and antinociceptive mechanisms. Temporal summation of pain (TSP), a gradual increment of the pain sensation elicited by repeated C-fiber–mediated stimuli which is evaluated as a measure of pronociceptive mechanisms, is enhanced in NSCNP patients(14,16,18). In addition, disruption of endogenous antinociception has also been found, such as the impairment of the so-termed Conditioned Pain Modulation (CPM)(15,16).

Although changes in central nervous system pain processing have been shown following manual therapy intervention, most studies have relied on static psychophysical measures (largely PPTs), and found reduction in local(19–21) and in some cases distal hyperalgesia(22,23). However, studies assessing the effects of manual therapy using dynamic psychophysical tests are scarce, and relatively little is known on the effects of manual therapy on central pain processing mechanisms. Dynamic psychophysical tests have been postulated to better assess central nervous system pain processing(24) since they evaluate central nervous system mechanisms rather than signs. Although a systematic review showed that physical therapy may reverse alterations in pain processing that accompany several musculoskeletal conditions(25), few studies have specifically addressed the effect of manual therapy, and only one considered NSCNP(26) treated by neurodynamic upper limb mobilizations, which found beneficial effects of therapy on CPM but not on TSP. Other studies in patients with NSCNP found no effects of treatment on central pain processing mechanisms(27,28), however none of them involved manual therapy.

For normalization of central pain processing to be considered as a potential mechanism of action of manual therapy, normalization should be associated with improvements in clinical outcome(29). The only studies that have so far addressed this issue have relied on PPTs as the sole measure of central pain processing, and failed to find associations between clinical outcome and changes in mechanical pain thresholds(30,31). No study has assessed the association between changes in dynamic measures of pain processing and clinical outcome following manual therapy.

The present study aimed to determine whether manual therapy restores normal functioning of central pain processing mechanisms in patients with NSCNP. As a secondary aim, we sought to evaluate the relationship between clinical outcome and changes in central pain processing mechanisms following treatment with manual therapy.

## Methods

A single-center, prospective study was conducted at a primary care physiotherapy clinic in the Bizkaia region of Spain between March 2020 and July 2021. All patients provided written consent before data collection and their rights were protected. The study was approved by the institutional review board at the University of the Basque Country–UPV/EHU (Ethical approval reference: M10_2018_160MR1_ZABALA MATA) and registered before study commencement (ClinicalTrials.gov record number: ACTRN12620000163909).

### Participants

In a two-sided contrast, with an alpha risk of 5%, a statistical power of 80%, and assuming maximum indeterminacy, 63 subjects were required to detect a maximum difference of 10% in TSP and CPM measures. People seeking treatment for NSNP at a primary care physiotherapy clinic were invited to participate. Inclusion and exclusion criteria for participants are shown in **Table 1**.

**Table 1.**
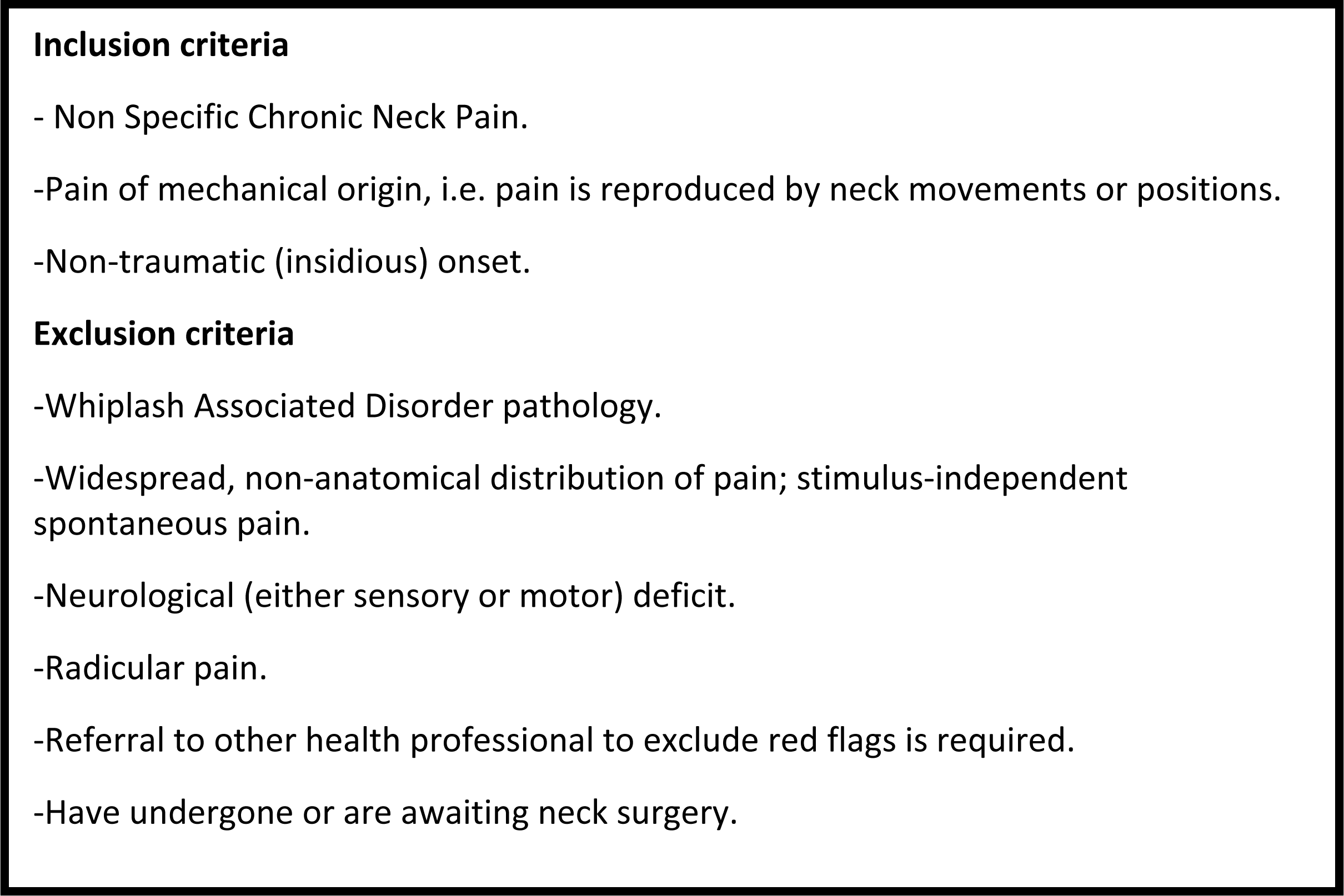
Inclusion/exclusion criteria.

### Clinical Assessment

Age, sex, height, and weight were recorded from participants, and patients completed the Neck Disability Index (NDI), the Pain Catastrophizing Scale (PCS) and The Tampa Scale of Kinesophobia (TSK) questionnaires. The NDI is a widely used, self-administered questionnaire for assessing cervical disability. The questionnaire consists of 10 items on activities of daily living, and each item is scored from 0 to 5, where higher scores indicate greater disability(32). The PCS is a 13-item questionnaire that measures catastrophic thoughts and feelings about pain(33). Total scores range from 0 to 52, and higher scores indicate higher levels of pain-related catastrophizing. Pain-related fear of movement was assessed using the 11-item TSK; scores on each item range from 1 to 4, where higher scores are indicative of greater fear(34).

Maximum and average intensities of pain experienced over the last 24 hours, and pain experienced during neck movements (flexion, extension, right and left rotation and side flexion) were recorded using a 0-10 numeric rating scale anchored with 0= no pain at all to 10= worst pain imaginable. Patients were also asked to complete the Patient Specific Functional Scale (PSFS)(35), a self-reported measure of perceived level of disability on specific items relevant for them.

In addition, patients were asked to rate their perceived treatment effect using the Global Rating of Change Scale (GROC). The GROC is a 15-point scale where clinical change is rated from -7 (a very great deal worse), through 0 (no change), to +7 (a great deal better)(36).

All clinical measures except GROC (recorded only post-treatment) were obtained in single sessions both at baseline and after treatment completion. A maximum of 24 hours elapsed both from the first clinical assessment session to treatment initiation, and from the last treatment session to the second clinical assessment.

### Psychophysical assessment

Pressure Pain Thresholds (PPT), defined as the minimum pressure at which pressure sensation becomes a painful sensation(37), were measured at several locations using a digital hand-held algometer with a 1- cm^2^-diameter rubber tip (Fisher, Pain Diagnostics and Thermography Inc, Great Neck, NY, USA). For local assessment of pain sensitivity, PPTs were measured bilaterally at the angle of the upper trapezius fibers, 5 and 8 cm above and medial to the superior angle of the scapula, and remote sensitivity was assessed on the tibialis anterior at a location 2.5 cm lateral and 5 cm inferior to the anterior tibial tuberosity. Subjects were instructed to report their first perceived pain sensation during an incremental pressure application at 1 kg/ sec. The same procedure was repeated three times, 1 min apart, and the average of three measurements was used for analysis. Patients were familiarized with the measurement protocol prior to the actual measurements. This procedure has shown high reliability in neck pain patients(38).

For assessing TSP, patients were seated in a quiet room with their hand rested on a table (same side as neck pain, or the side of most painful neck pain in patients with bilateral pain) and two adhesive Ag/AgCl electrodes were placed on the hand dorsum, 2 cm apart. Electrical stimuli consisting of brief bursts of five, 1 ms-long positive-square pulses, were generated by a constant current electrical stimulator (DS7; Digitimer Ltd, Welwyn Garden City, UK) and delivered at 200 Hz(39), which were perceived by the participant as single stimuli. Electrical pain thresholds were first determined using the increasing and decreasing staircase method with 0.2 mA stimulus increments/decrements. The electrical pain threshold was defined as the minimum current intensity evoking a sensation rated as painful in an incremental series or the current intensity no longer evoking pain in a decremental series, and the final value was recorded as the average of three consecutive incremental and three decremental measures. For assessing TSP, a single stimulus was administered at 1.2 times the electrical pain threshold intensity, and the participant was asked to rate the evoked pain sensation on a 0–100 numeric scale where 0 denotes no pain at all and 100 indicates the worst pain imaginable. Two minutes thereafter, 5 consecutive stimuli of the same current intensity were delivered at a frequency of 2 Hz (2.5-millisecond total stimulus duration), and the participant was asked to rate the pain sensation evoked by the stimulus perceived as the most painful. The ratio of the second rating to the first was used as the TSP measure(40). A higher ratio was indicative of greater TSP. This protocol has been previously used(41) and is based on well-known parameters for evaluating TSP(39).

For CPM assessment, PPT was measured first on the trapezius muscle as above, and the participant was then asked to immerse his/her contralateral foot in cold water (kept at 10° C) for 2 minutes or until pain became unbearable. Immediately thereafter, the PPT was measured again at the same location. The CPM response was obtained by subtracting the second measure from the first(42). A greater value was indicative of higher endogenous pain inhibition. This procedure has demonstrated good to very good reliability(43).

### Intervention

Patients received weekly, 45-minute sessions of manual therapy for 4 weeks. Treatments consisted of articular passive mobilizations, soft tissue mobilization, and trigger point treatment performed by the clinician following clinical reasoning. Passive mobilization treatment consisted of passive, low-speed movements performed on hypomobile and pain-reproducing spinal segments in the cervical and thoracic spine(36,44), including segmental translations and physiological movements both through and at the end of the range of movement(45). Soft tissue mobilization (gentle longitudinal and transverse stroking) of neck muscles was administered in order to improve connective tissue function and reduce myofascial pain. This was accompanied by a trigger point technique on neck muscles were appropriate(46). All treatments were administered by a physiotherapist with postgraduate training and 15 years of experience in musculoskeletal physiotherapy.

Two sets of generalized linear mixed model (GLMM), with Gaussian response and the identity link (i.e. equivalent to a linear regression), were used to assess the effect of the intervention on clinical, psychological and psychophysical measures, and the association between treatment-induced psychophysical changes and clinical and psychological outcomes. Analyses were controlled for sex, age, BMI, baseline value of the variables of interest, and individual heterogeneity. Individual heterogeneity, controlled for including a random effect, collects unobserved invariant variables over time that are specific to each individual participant, i.e. residual confounding. Given the complexity of the models, we performed inferences using a Bayesian framework. In particular, we followed the Integrated Nested Laplace Approximation (INLA) approach(47,48). In addition to the coefficient estimators and their 95% credibility intervals, the probability of the coefficient estimator (an absolute value being more than 1 (Prob(|estimator|)>1), Prob, was also computed (note that this is unilateral and may not coincide with the credibility interval). Unlike the p-value in a frequentist approach, this probability allows us to make inferences about associations between dependent and independent variables. For the sake of simplicity, Prob values exceeding 0.95 are equivalent to p<.05 in a non-Bayesian context. All analyses were conducted using the open access software *R* (version 4.2.2)(49) available through the INLA package(47,48,50)

## Results

Sixty-three participants took part in the study between 03/03/2020 and 21/07/2021. Demographics and baseline clinical characteristics are shown in **Table 2**. All participants attended the scheduled therapy sessions and completed the treatment, and there were no drop outs (Fig 1).

**Fig 1.**
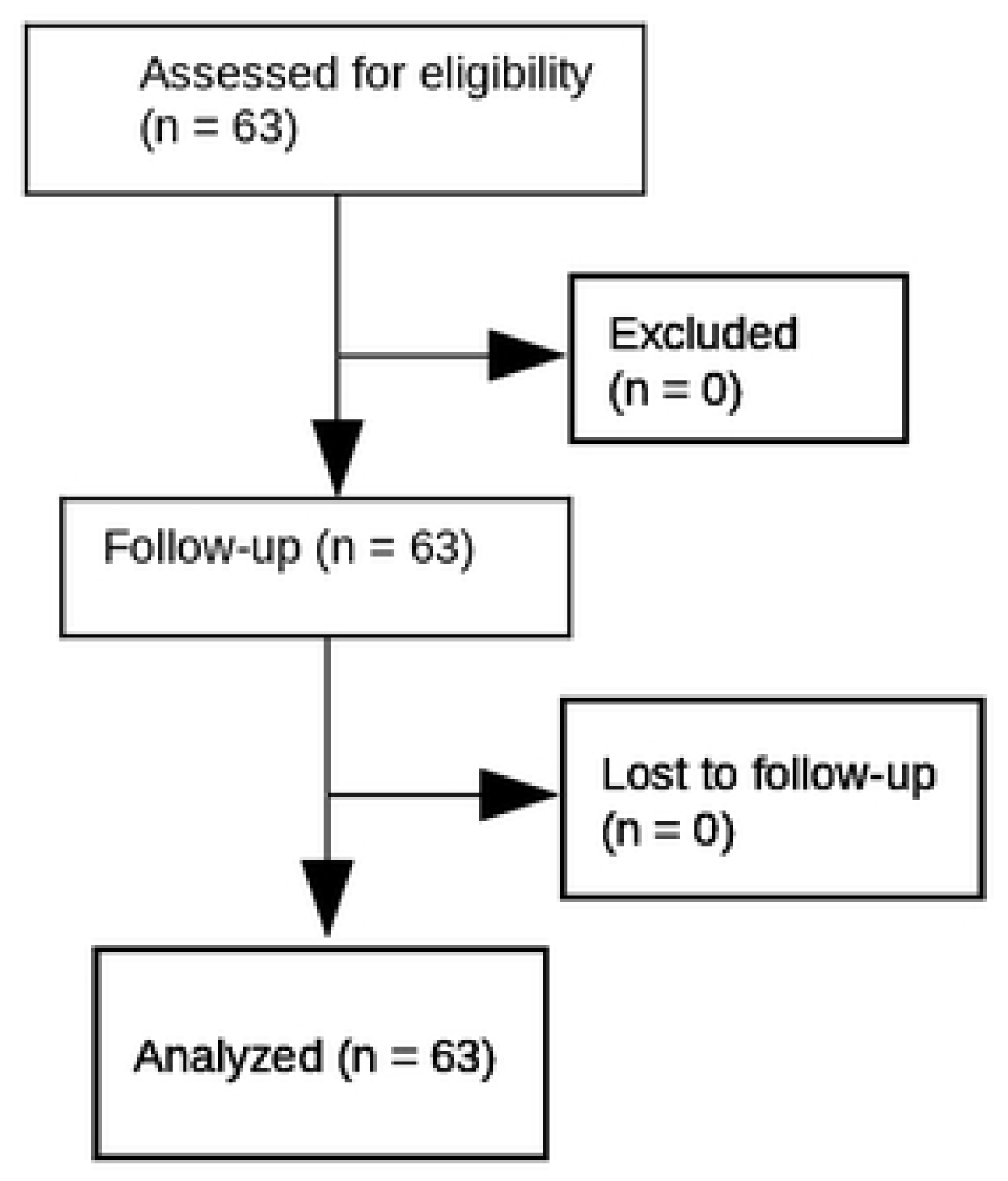
CONSORT Flow Diagram.

**Table 2.** Demographics and clinical Characteristics. Values are mean (SD) or number of cases.

|  |  |
| --- | --- |
| N | 63 |
| Sex (f/m) | 50/13 |
| Age (y) | 45.8 (14.3) |
| BMI | 23.5 (3.2) |
| Neck pain duration (y) | 6.7 (5.2) |
| Average pain 24 hours (0–10) | 4.72 (1.83) |
| Maximum pain 24 hours (0–10) | 6.26 (1.82) |
| NDI (0–50) | 11.56 (5.23) |
| PCS (0–52) | 15.38 (9.28) |
| TSK (0–44) | 23.99 (7.29) |
| PSFS (0–10) | 4.24 (1.93) |
| Pain on movement | 3.48 (2.03) |

Abbreviations: BMI, body mass index; NDI, neck disability index; PCS, pain catastrophizing scale; TSK, Tampa scale kinesophobia; PSFS, Patient specific functional scale; TSP, temporal summation pain; CPM, conditioned pain modulation.

### Changes in pain processing, and clinical and psychological outcomes following the intervention

Patients showed an improvement in central pain processing following manual therapy. Namely, the intervention both attenuated TSP response (Coefficient: -0.63; 95% credibility interval = -0.82 to -0.43; P= 1.00) and improved conditioned modulation of pain (Coefficient: 0.89; 95% credibility interval = 0.14 to 1.65; P= .99). In addition, manual therapy increased PPT on the trapezius muscle on the side of neck pain (Coefficient: 0.22; 95% credibility interval = 0.03 to 0.42; P= .98), but not on the contralateral trapezius (Coefficient: 0.01; 95% credibility interval = -0.19 to 0.21; P= .54) or the tibialis anterior muscle (ipsilateral Coefficient: -0.03; 95% credibility interval = -0.29 to 0.22; P= .59 on the side of neck pain. Contralateralcoefficient: 0.01; 95% credibility interval = -0.26 to 0.29; P= .54 contralaterally) (Table 3).

**Table 3.** Effect of intervention on clinical, psychological and psychophysical variables. Model adjusted for sex, age, BMI, individual heterogeneity (random effect) and baseline value of variable.

| Variables | Coefficient | 95% Credibility Interval | P |
| --- | --- | --- | --- |
| NDI (50) | -2.23 | 1.29 to 3.17 | 0.99 * |
| PCS (52) | -7.43 | -9.01 to -5.85 | 1.00 * |
| TSK (44) | -4.07 | -5.25 to -2.88 | 1.00 * |
| PSFS_average | 2.58 | 1.89 to 3.26 | 1.00 * |
| Maximum pain 24h | -3.05 | -3.53 to -2.58 | 1.00 * |
| Average pain 24h | -2.52 | -2.91 to -2.13 | 1.00 * |
| Pain on movement | -2.19 | -2.51 to -1.87 | 1.00 * |
| PPT ipsilateral trapezius | 0.22 | 0.03 to 0.42 | 0.98 * |
| PPT contralateral trapezius | 0.01 | -0.19 to 0.21 | 0.54 |
| PPT ipsilateral TA | -0.02 | -0.28 to 0.23 | 0.59 |
| PPT contralateral TA | 0.01 | -0.25 to 0.29 | 0.54 |
| TSP change (ratio) | -0.63 | -0.82 to -0.43 | 1.00 * |
| CPM change (absolute) | 0.89 | 0.14 to 1.65 | 0.99 * |

Abbreviations: BMI, body mass index; NDI, neck disability index; PCS, pain catastrophizing scale; TSK, Tampa scale kinesophobia; PSFS, Patient specific functional scale; PPT, pressure pain threshold; TA, tibialis anterior; TSP, temporal summation pain; CPM, conditioned pain modulation.
\* Statistically significant change.

Clinical pain was also ameliorated following manual therapy, as shown by the reduction in average pain ratings at 24 hours (Coefficient: -2.52; 95% credibility interval = -2.92 to -2.13; P= 1.00), maximal pain ratings at 24 hours (Coefficient: -3.07; 95% credibility interval = -3.54 to -2.59; P= 1.00) and pain ratings during neck movements (Coefficient: -2.19; 95% credibility interval = -2.51 to -1.87; P= 1.00) (Table 3). The majority of patients reported feeling “a very great deal better” or “a great deal better” (21% and 33% respectively) following treatment, 16% reported feeling “quite a bit better”, 12% “moderately better”, 5% “somewhat better”, 5% “a little bit better”, 6% “a tiny bit better” and 2% “about the same”. Favorable changes in functional and psychological status were also noted, as shown by statistically significant improvements in measures of disability (Coefficient: 2.24; 95% credibility interval = 1.28 to 3.19; P= .99), function (Coefficient: 2.58; 95% credibility interval = 1.89 to 3.26; P= 1.00), fear of movement (Coefficient: -4.04; 95% credibility interval = -5.24 to -2.85; P= 1.00) and catastrophization (Coefficient: -7.41; 95% credibility interval = -9.00 to -5.82; P= 1.00) (**Table 3**).

### Association between changes in central pain processing and clinical and psychological outcomes

Improvements in the functioning of central pain modulatory mechanisms (CPM and TSP) following intervention were found to be very weakly associated with only few measures of clinical and psychological outcome as shown in **Table 4**. Improvement in the CPM response was found to be negatively correlated with changes in PSFS (Coefficient: -0.65; 95% credibility interval = -1.22 to -0.07; P= 0.98), and attenuation of TSP was found to be associated with a greater improvement in pain during movement (Coefficient: 0.42; 95% credibility interval = 0.10 to 0.74; P= 0.99).

**Table 4.**
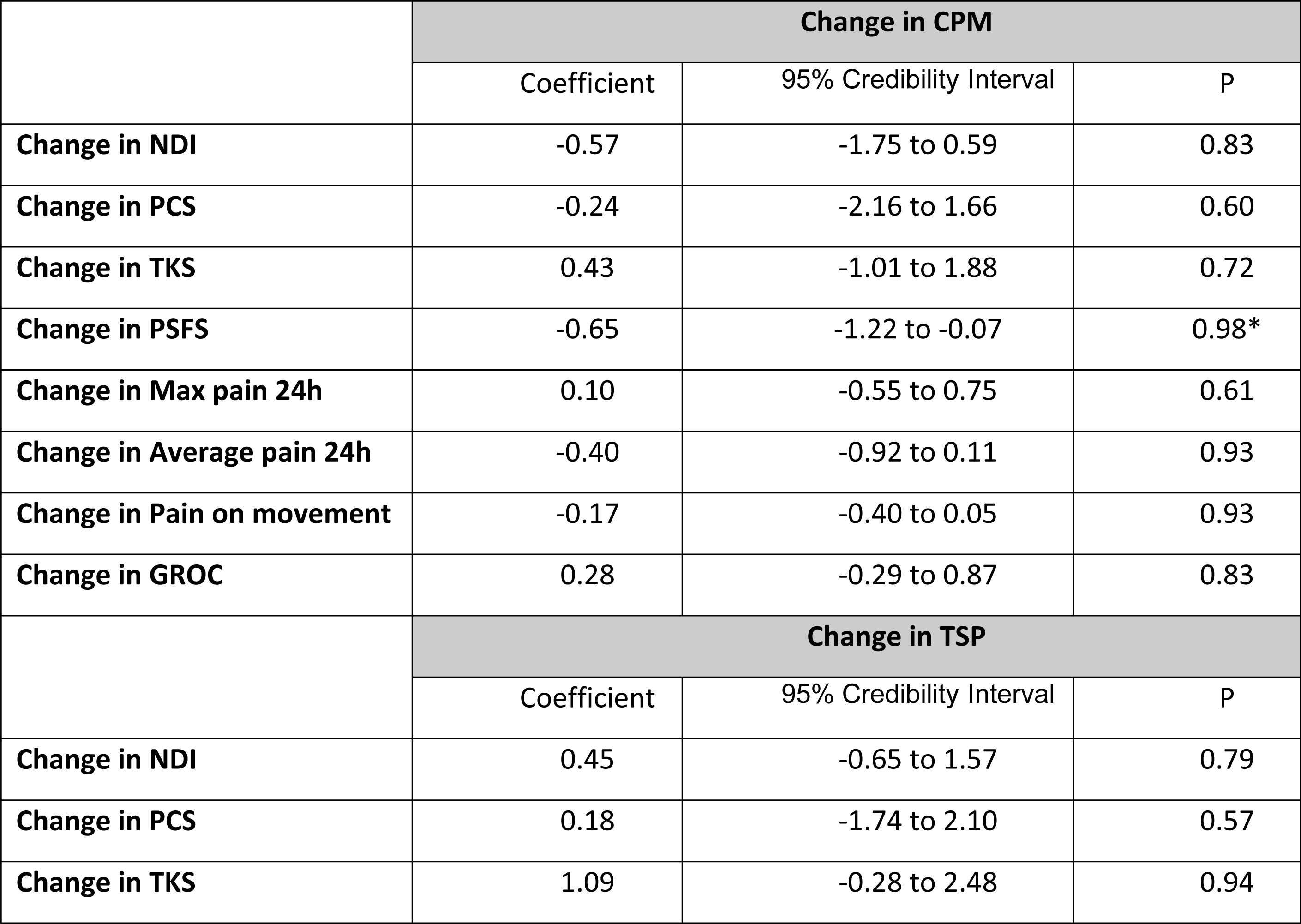
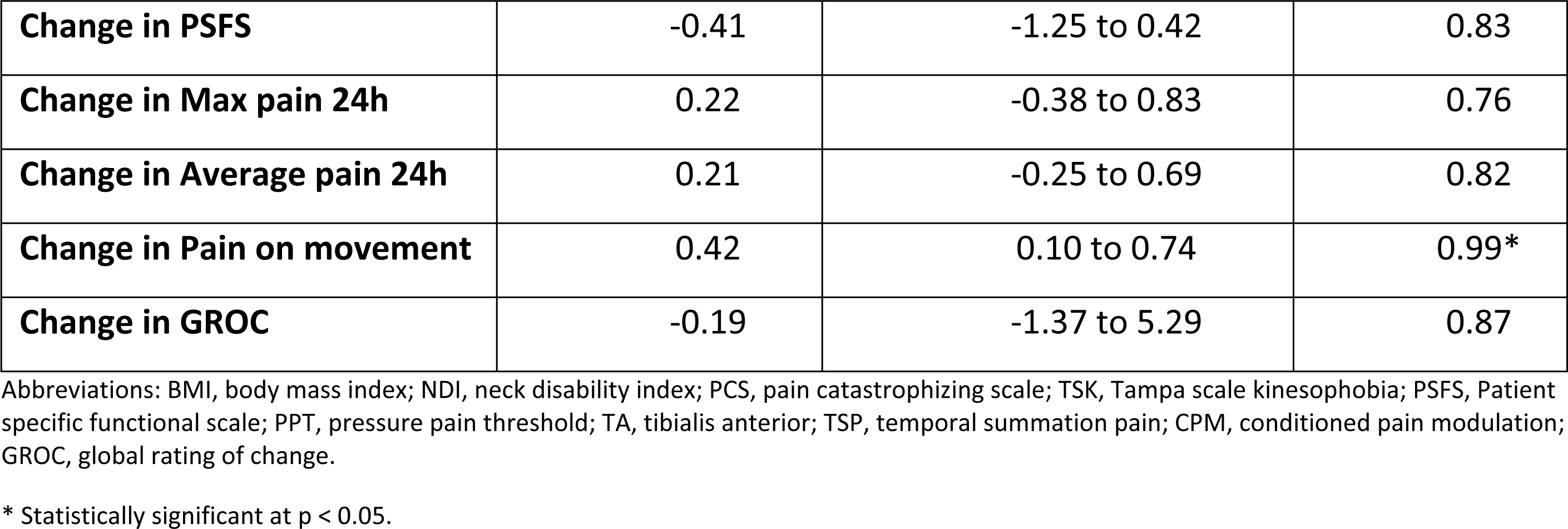
Association between changes in central pain processing mechanisms and clinical/psychological variables. Model adjusted for sex, age, BMI and individual heterogeneity (random effect).

## Discussion

The present work provides novel evidence of restoration of normal central pain processing following manual therapy in NSCNP patients, as shown by TSP, CPM and PPT values returning to normal levels (i.e. comparable to normative data collected in our lab(16)). In addition, clinical pain was ameliorated, and both functional and psychological measures were improved.

Significant attenuation of TSP was noted following manual therapy. A normalizing effect of manual therapy on temporal summation has been previously reported both in healthy volunteers(51) and pain conditions such as low back pain(52) and carpal tunnel syndrome(53). In contrast, previous studies evaluating physical therapy outcome in patients with NSCNP have failed to report changes in TSP following treatment with a variety of interventions including virtual reality(28), cervical therapeutic exercise(28,54) and a combined protocol of electrotherapy and cervical therapeutic exercise^28^. It is possible that TSP may preferentially or to a greater extent be affected by manual therapy in patients with NSCNP, but not by other treatment modalities. Alternatively, methodological differences related to the ability of the employed stimuli to activate afferent nociceptive pathways may have also contributed to this disparity of results, since the mechanical stimuli used to measure TSP in previous studies (weighted pinprick stimuli(27), or pressure exerted by either a cuff (54) or an algometer(28)) stimulate superficial and deep nociceptors whereas the electrical stimuli used in the current study may directly recruit afferent C-fibers(55). Moreover, electrical stimulation has been shown to be more effective at producing cortical somatosensory activation(56).

We found that manual therapy also restored CPM response in our cohort. This finding is consistent with a previous study where neurodynamic treatment, i.e. a form of manual therapy, improved CPM in a similar patient population^27^. Indeed, several studies have shown a general normalizing effect of treatment on CPM response in NSCNP(26,54,57) and in other pain conditions(58,59) regardless of the therapeutic intervention. It thus appears that the normalizing effects of interventions on CPM are frequent, and less dependent on the type of treatment or target population.

The relationship between an increased TSP response and the level of perceived pain in the NSCNP population is not entirely clear. Although a systematic review with meta-analysis did find some association in a population of back pain patients(60), two recent case-control studies studying NSCNP failed to do so(16,61). In light of the fact that the expression of wind up at spinal cord neurons may be genetically encoded(62), an enhanced TSP may be viewed as an indicator of higher propensity to developing pain hypersensitivity. In support of this notion, a number of studies have shown TSP to be a predictor of pain prospectively(63–65). From a neurophysiological standpoint, temporal summation is considered as one of the initiating neuroplastic mechanisms of central sensitization(66,67). On the other hand, wind up as the correlate of TSP in animal models has also been found to be profoundly influenced by descending supraspinal modulation(68–70). It thus seems reasonable to assume that the reducing effects of manual therapy on TSP as shown here may, at least in part, be produced by recruiting central pain modulatory mechanisms. This view is further strengthened by the enhancing effect of manual therapy on the CPM response in patients with NSCNP as shown here. The CPM paradigm is an experimental model to assess the functional state of the so-termed Diffuse Noxious Inhibitory Controls, a widespread modulatory mechanism arising from the brainstem and operating on second order wide dynamic range neurons in the spinal dorsal horn via the spinal dorsolateral funiculi(71). An enhanced CPM is considered to reflect greater efficacy of endogenous analgesia mechanisms and thus a more favorable position to control central excitation induced by incoming peripheral nociceptive input(72).

Whilst pain processing and clinical status were both improved following therapy, we found no distinct relationship between the two types of outcomes. Therefore, our present results provide no support for normalization of central pain processing as the sole or main mechanism of action of manual therapy in NSCNP. However, it is likely that the nature of our sample may have contributed, at least in part, to this lack of association. Although central pain processing mechanisms at baseline were altered as compared to controls(16), alterations were indeed modest and patients presented only mild baseline disability(73), therefore any clinical improvement could be expected to be accompanied by only limited beneficial effects on pain processing.

The physiological mechanisms underlying the clinical effect of manual therapy on NSCNP remain unclear. Collectively, however, our present observations of the beneficial effects on TSP and CPM responses support the notion that manual therapy may operate, to some extent, by influencing central pain processing to ameliorate pain and improve clinical status. Nonetheless, the fact that no clear association was observed between restoration of normal central pain processing and clinical outcome suggests a plurality of underlying mechanisms that may also likely involve biomechanical, physiological and psychological changes. More studies are needed to determine specifically which mechanisms of action influence the clinical improvement of NSCNP with manual therapy.

## Limitations

Since no control group was used, changes in both clinical and pain measures noted here cannot unequivocally be attributed to the intervention. However, no study to date has reported spontaneous normalization of central pain processing, and the fact that clinical improvement following the intervention was achieved after three months of persisting clinical manifestations renders an alternative explanation less likely.

The present study only measured outcomes in the short term, and thus whether the observed changes are long lasting was not determined.

## Data Availability

All relevant data are within the manuscript and its Supporting Information files.

## Acknowledgments

I would like to express my gratitude to Hiru Fisioterapia for generously offering us their facility as a research location.

